# Whom Does Algorithmic Risk Stratification Miss? A Fairness Audit of Machine Learning Targeting for Concurrent Maternal–Child Double Burden of Malnutrition Across 30 Low- and Middle-Income Countries

**DOI:** 10.64898/2026.04.28.26352000

**Authors:** Xiaoyu Wu, Benqing Zheng

## Abstract

**Background:** Concurrent maternal–child double burden of malnutrition (DBM) affects a growing share of mother–child dyads in low- and middle-income countries (LMICs). Nutrition programmes often use maternal education as an eligibility proxy, but whether algorithmic alternatives would do better—and at what equity cost—has not been directly tested. We evaluated whether machine learning (ML)-based targeting for two concurrent DBM subtypes—overweight mother with stunted or wasted child (Subtype A) and underweight mother with stunted or wasted child (Subtype B)—improves recall over a proxy-based rule while preserving fairness across social strata.

**Methods:** We pooled Phase 7–8 Demographic and Health Surveys from 30 LMICs (181,636 mother–child dyads). We first estimated subtype-specific social gradients with multilevel logistic regression. We then trained xgboost prediction models with strict label-leakage safeguards and leave-one-country-out cross-validation, and compared ML-based targeting against random and education-based rules at 10%, 20%, and 30% budget constraints. Fairness was audited along six social strata using equalized-odds, demographic-parity, calibration, and predictive-value gaps. A full-India sensitivity analysis (354,691 dyads) assessed robustness to down-sampling.

**Findings:** Overall weighted any-DBM prevalence was 12.52% (Subtype A: 8.21%; Subtype B: 4.31%). Subtype A showed an inverted-U gradient on wealth (adjusted odds ratio peak 1.22 at Richer versus Poorest) and maternal education (peak 1.23 at Primary versus None); Subtype B declined monotonically (wealth: 0.25 at Richest; higher education: 0.49). Mean leave-one-country-out area under the curve was 0.615 for Subtype A and 0.652 for Subtype B. At a 20% budget, ML captured 35.3% of Subtype A cases versus 18.4% for education-based targeting (+92%); for Subtype B the corresponding values were 37.5% and 32.1% (+17%). Equalized-odds gaps reached 0.57 on country income and 0.59 on maternal education; true-positive rates were lowest in the highest-wealth and highest-education strata. Results were stable under the full-India sensitivity analysis.

**Conclusions:** ML is useful principally for Subtype A, where the education proxy is no better than random. For Subtype B it mostly changes who gets reached rather than how many, which is a policy choice rather than an accuracy upgrade. The households the algorithm most often misses are not the poor but the rare positives in high-resource strata, which is what a fixed-budget rule ranking on heterogeneous base rates will do. Programmes should decide whether their priority is total capture or the distribution of capture before adopting such a rule.

**Author Summary:** In many low- and middle-income countries, mothers who are overweight often live in the same household as children who are too short or too thin for their age. Nutrition programmes that try to reach such families have limited resources, so they must choose which households to prioritise. Most programmes use maternal education level as a rough filter, but whether this is actually a good way to find affected families has rarely been tested.

We used surveys of 181,636 mother–child pairs from 30 low- and middle-income countries to compare three ways of identifying at-risk households: random selection, selection by low maternal education, and selection by a machine-learning model. Machine learning was much better at finding families where an overweight mother lives with an undernourished child—nearly doubling the capture rate compared with the education rule. For a different combination (underweight mother with an undernourished child), machine learning did not clearly outperform education on total recall; instead it reached different households, mostly shifting attention toward the rural poor.

An unexpected finding was that the households the algorithm was most likely to miss were not the poor ones, but the wealthier and better-educated ones, where this type of malnutrition is rarer. This is not bias against the poor—it is what happens when any ranking rule operates under a fixed budget. Programmes that want to reach everyone at risk, regardless of how rare risk is in a given group, may need more than one rule.

## 1. Introduction

Child undernutrition remains a defining global-health challenge. In 2024, 150.2 million children under five years—nearly one in four globally—were stunted, 42.8 million (6.6%) were wasted, and 35.5 million (5.5%) were overweight [1]. What these aggregate figures do not show is that tens of millions of LMIC households now experience maternal overweight and child undernutrition at the same time [2,3]. Concurrent maternal–child double burden of malnutrition (DBM) is no longer confined to countries deep in nutrition transition; it has become central to the second Sustainable Development Goal [4–7].

What we still do not know is how to screen for it. Chen and colleagues [8], in a pooled 45-country DHS analysis of 423,340 mother–child pairs, showed that maternal education was inversely associated with overall DBM (odds ratio [OR] 0.71 for tertiary versus no education) but positively associated with subtypes involving overnutrition—so the direction of the education effect depends entirely on which subtype is at issue. Seferidi and colleagues [9] added a temporal dimension: within-country probability of the stunted-child/overweight-mother combination has risen at 1.04 times per year across 55 LMICs since 1992, driven more by social than by macroeconomic globalisation. A longer tradition has documented that the obesity–socioeconomic-status relationship in LMICs is non-monotonic [10–12], and regional work has mapped the local texture of these gradients in South and Southeast Asia [13] and sub-Saharan Africa [14], while related work has examined income and education disparities in childhood malnutrition more broadly [15]. These are aetiological studies. A risk factor, however, is not a decision rule. Whether maternal education—a simple, low-cost proxy often available in field surveys—is good enough as an eligibility criterion has not been tested against any algorithmic alternative.

Machine learning is one obvious candidate. Aiken and colleagues [16] used phone and satellite data to redirect humanitarian cash transfers in Togo toward households that standard geographic rules had missed, and Smythe and Blumenstock [17] reported related gains for geographic microtargeting of social assistance using high-resolution poverty maps. More broadly, development economists have long debated when proxy-means testing improves targeting and when it excludes intended beneficiaries [18,19]. But ML has its own problem. Obermeyer and colleagues [20] showed how an ostensibly neutral US healthcare algorithm systematically underserved Black patients because its training label encoded racialised access patterns, and a subsequent methodological literature has proven that no algorithm can simultaneously satisfy calibration, equalized odds, and demographic parity when group prevalences differ [21–23]. Fairness is now a scientific precondition for deployment, not a post-hoc ethical check [24–27]. Yet a recent meta-analysis of 11 DHS-based ML studies of child malnutrition found that fairness analyses have been essentially absent from this literature [28].

We use DHS Phase 7–8 data from 30 LMICs to ask three questions. Do Subtype A (overweight mother with stunted or wasted child) and Subtype B (underweight mother with stunted or wasted child) follow distinct enough social gradients that a single targeting rule would work differently on the two? Does an ML-based rule beat the common education proxy, and by how much for each subtype? And if it does, does that gain come at an equity cost—and which groups pay it?

## 2. Materials and methods

### 2.1. Data source and study population

We analysed the Demographic and Health Surveys (DHS), a programme that has conducted more than 400 nationally representative household surveys in over 90 LMICs since 1984 under standardised protocols [29,30]. We included the most recent DHS Phase 7 or Phase 8 survey for each country that collected complete mother-and-child anthropometry for women aged 15–49 years and their most recent live birth aged 0–59 months. Eligibility required: (i) valid maternal height and weight measurements; (ii) valid child height-for-age, weight-for-age, and weight-for-height z-scores computed against the WHO Child Growth Standards [31], with data quality assessed following Assaf and colleagues [32]; (iii) observed household wealth quintile (v190 [33,34]), maternal completed education (v106), and urban/rural residence. Three otherwise eligible surveys (Colombia 2015; Indonesia 2017; the Philippines 2022) were excluded because the most recent rounds did not collect anthropometric measurements, and Kazakhstan, Mexico, and Uzbekistan were excluded for the same reason. Ethiopia 2019 and Senegal 2019 were excluded because maternal height and weight variables (v437, v438) were entirely missing from the public DHS recodes available to us, resulting in zero analytic-ready dyads after anthropometric harmonisation. The final analytic pool comprised 181,636 mother–child dyads from 30 countries. Because India’s unrestricted DHS contribution (197,932 dyads) would have dominated model estimation, we down-sampled India by primary sampling unit (PSU) to 24,870 dyads (13.7% of the analytic pool); a full-India sensitivity analysis (n = 354,691) is reported in Section 3.6.

### 2.2. Outcome definitions

Following the Lancet DBM series’ within-dyad framing [2,3] we defined two mutually exclusive concurrent DBM subtypes. Subtype A comprised dyads in which the mother was overweight (body-mass index [BMI] ≥ 25 kg/m^2^) and the child was stunted (height-for-age z-score [HAZ] < −2) or wasted (weight-for-height z-score [WHZ] < −2). Subtype B comprised dyads in which the mother was underweight (BMI < 18.5 kg/m^2^) and the child was stunted or wasted. Because the two maternal BMI conditions are disjoint by construction, a dyad cannot belong to both. Any-DBM was the union of A and B.

### 2.3. Exposures and covariates

Individual-level variables comprised maternal age in years (grouped 15–24, 25–34, 35–49), completed maternal education (none, primary, secondary, higher), wealth quintile (Poorest to Richest), place of residence (urban, rural), child sex, and child age category (0–11, 12–23, 24–35, 36–47, 48–59 months). Country-level variables comprised the World Bank income-group classification contemporaneous with the survey year (low, lower-middle, upper-middle) and a six-level region indicator (South Asia; Southeast Asia; Central Asia; sub-Saharan Africa; Latin America and Caribbean). Complete case analysis was used; fewer than 0.2% of the analytic pool were excluded for covariate missingness.

### 2.4. Multilevel logistic regression

We estimated two-level random-intercept logistic models with dyads nested in countries, separately for Subtype A and Subtype B, using the R package lme4 with bobyqa optimisation and Laplace approximation. Fixed effects included maternal age group, maternal education, wealth quintile, residence, child sex, child age group, and country income group; the country-level random intercept captured unexplained between-country heterogeneity. Because our primary interest was in between-country heterogeneity and subtype-specific social gradients, we specified country-level random intercepts and used DHS weights as an approximation to the complex sampling design, rather than estimating a full three-level design-based model [30]. The intra-class correlation coefficient was computed on the latent-response scale. DHS sampling weights (v005/10^6^) were incorporated as probability weights in model fitting. Reference categories were Poorest, no education, urban, male, 25–34 years, 24–35 months, and low-income country.

### 2.5. Machine learning model and leave-one-country-out cross-validation

We trained xgboost gradient-boosted classifiers [35] separately for Subtype A and Subtype B. The feature set deliberately excluded all anthropometric variables and their derivatives (maternal BMI and its components, all child z-scores and binary indicators, and maternal over-/underweight flags) to prevent label leakage; the remaining features comprised maternal age, maternal education level, wealth quintile, residence, child sex, child age, and dummy-coded country income group. A label-leakage audit fitting xgboost on only the excluded anthropometric variables verified that these variables alone were near-perfectly informative of the outcome (area under the curve > 0.99 for each outcome), confirming that their exclusion from the main feature set was necessary rather than conservative. We used leave-one-country-out cross-validation with 30 folds for external validation—an evaluation design closely matching the most demanding deployment scenario (transfer to a new country)—an inner three-fold cross-validation on the training countries for hyperparameter tuning across an eight-point grid (tree depth 4 or 6, learning rate 0.05 or 0.1, minimum child weight 1 or 5), and early stopping at 20 rounds on the validation area under the curve. DHS sampling weights (v005/10^6^) were supplied as per-instance weights via the weight argument of xgb.DMatrix; xgboost treats these as instance weights rather than frequency weights (they do not replicate observations but rescale each dyad’s contribution to the gradient), and the same weights were used for survey-representative performance estimation. Performance was summarised by area under the receiver-operating-characteristic curve (AUC), Brier score, and calibration-in-the-large, each computed per fold, following TRIPOD+AI reporting guidance [26].

### 2.6. Targeting comparison under budget constraints

We compared three targeting rules under fixed population-level budget constraints of 10%, 20%, and 30%, reflecting feasible intervention coverage in LMIC nutrition programmes [36]. The rules were: (i) random assignment; (ii) education-based, in which dyads with the lowest maternal education were selected first with ties broken by a small uniform perturbation; and (iii) ML-based, in which dyads were ranked by the held-out xgboost-predicted probability. Education-based targeting served as the primary policy baseline because it is interpretable, low-cost, and already widely used as an eligibility proxy in LMIC cash-transfer and nutrition programmes—its value is programmatic rather than statistical, and comparing ML against it addresses the policy-relevant question of whether a more complex rule is worth adopting [18,19]. The 20% budget served as the primary operating point because it represents a realistic constrained-coverage scenario rather than universal screening; we additionally report 10% and 30% to test robustness to this choice. For each rule, subtype, and budget we computed weighted recall (the proportion of true DBM cases captured) and precision (positive predictive value). Ninety-five per cent confidence intervals were obtained by country-clustered bootstrap with 500 resamples.

### 2.7. Fairness audit

We audited the ML-based rule at the 20% budget threshold across six social strata—wealth quintile, residence, maternal education, country income group, child sex, and maternal age group. Four complementary metrics were reported per axis: demographic-parity gap (the range of the proportion receiving a positive prediction), equalized-odds gap (the maximum of within-axis true-positive-rate and false-positive-rate ranges), calibration gap (the range of the mean difference between predicted probability and observed prevalence), and predictive-value gap (the range of positive and negative predictive values). These four metrics are mathematically incompatible in general—Kleinberg and colleagues [23] and Chouldechova [22] formalise the impossibility theorem that calibration and equalized error rates cannot simultaneously be satisfied when group prevalences differ—so we pre-specified equalized odds as the primary metric because a budget-constrained targeting rule operates as a hard classifier and the principal harm of interest is missed cases (false negatives) rather than mis-scored probabilities [21,24]. For each axis we identified the worst-off group (lowest true-positive rate). A zero-sum decomposition quantified how many additional true positives the ML rule captured, relative to the education-based rule, within each wealth-by-residence stratum.

### 2.8. Sensitivity analyses and reporting standards

Three sensitivity analyses were undertaken. First, the xgboost leave-one-country-out pipeline was re-run on the full-India sample (354,691 dyads) without PSU down-sampling. Second, Benjamini– Hochberg false-discovery-rate correction at q = 0.05 was applied across the 12 subtype-by-axis equalized-odds gap tests [38]. Third, alternative thresholds at 10% and 30% budgets were used to test robustness of the fairness audit. Reporting follows STROBE for observational components [37] and TRIPOD+AI for the prediction and fairness components [26]. Analyses used R 4.3 with lme4, survey, xgboost, and pROC; set.seed(20260421) fixed the India PSU sampling and set.seed(20260422) fixed xgboost training and bootstrap resampling to ensure reproducibility. The complete analysis pipeline is permanently archived at Zenodo (https://doi.org/10.5281/zenodo.19731115).

### 2.9. Ethical approval

This study analyses publicly available, de-identified secondary data provided by the DHS Program. No additional ethical approval was required; each original DHS survey received ethical clearance from the relevant national authorities and from the ICF Institutional Review Board.

## 3. Results

### 3.1. Sample characteristics and overall DBM prevalence

Overall, 12.52% of mother–child dyads experienced any form of DBM, with 8.21% classified as Subtype A (overweight mother with stunted or wasted child) and 4.31% as Subtype B (underweight mother with stunted or wasted child). Country-specific weighted any-DBM prevalence ranged from 6.1% in the Dominican Republic to 20.8% in Timor-Leste; Guatemala showed the highest Subtype A prevalence (19.7%) while Timor-Leste showed the highest Subtype B prevalence (13.9%) (Fig 1; Table 3). Our overall 12.52% weighted any-DBM prevalence is broadly consistent with, and slightly higher than, the 6.0% stunted-child/overweight-mother prevalence reported by Seferidi and colleagues [9] across 55 LMICs from 1992 to 2018, a discrepancy explained by our broader outcome definition (including underweight-mother subtypes and wasted children), the more recent survey window, and the secular annual rise that Seferidi and colleagues themselves documented.

**Fig 1.**
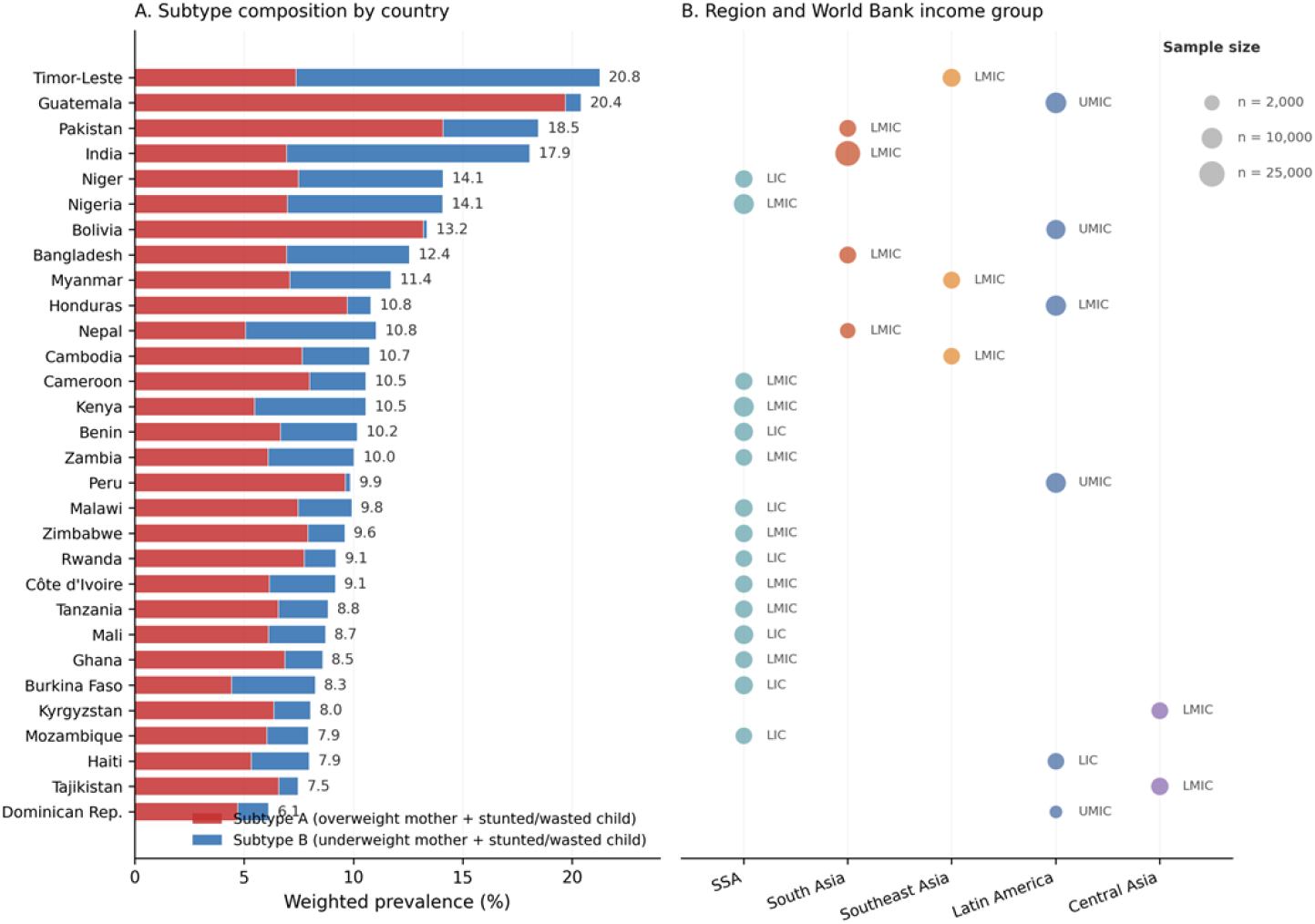
Weighted prevalence of concurrent maternal–child double burden of malnutrition across 30 LMICs, DHS Phase 7–8. Panel A: stacked bars showing Subtype A (red) and Subtype B (blue) composition, with total any-DBM percentage annotated to the right. Panel B: each country classified by region and World Bank income group; marker size is proportional to unweighted sample size.

The geography of subtype composition (Fig 1) is distinctive. Latin America and the Caribbean are dominated by Subtype A: in Guatemala and Bolivia, Subtype A accounts for essentially all DBM (Subtype B weighted prevalence <1%), while Peru and Honduras exhibit a less extreme but still strongly Subtype A–skewed pattern. South Asia shows the reverse: Timor-Leste, India, Bangladesh, and Nepal have Subtype B prevalence exceeding or matching Subtype A. Sub-Saharan African countries occupy a middle range, with balanced or slightly Subtype A–heavy distributions. Table 3 aggregates these country-level differences to the region × income-group level, following the stratification approach used by Seferidi and colleagues [9] and Popkin and colleagues [2]: Subtype A weighted prevalence rises from 6.6% in low-income countries to 7.3% in lower-middle-income to 14.4% in upper-middle-income, while Subtype B prevalence is highest in South Asia (9.5%) and Southeast Asia (8.3%), regions where maternal underweight remains demographically salient.

### 3.2. Inverted-U gradient for Subtype A and monotonic gradient for Subtype B

Multilevel logistic regression revealed distinct social gradients for the two subtypes (Fig 2; Table 1). For Subtype A, the wealth-quintile association was non-monotonic, with adjusted odds ratios rising from the Poorest reference through Poorer (1.10; 95% confidence interval [CI] 1.04–1.17), Middle (1.18; 95% CI 1.12–1.25), and peaking at the Richer quintile (1.22; 95% CI 1.14–1.30) before declining at the Richest quintile (1.17; 95% CI 1.09–1.26). The maternal-education gradient also displayed an inverted-U pattern, with Primary education carrying the highest adjusted risk (1.23; 95% CI 1.16–1.30 versus no education), followed by a monotonic decline through Secondary (0.89; 95% CI 0.83–0.94) and Higher education (0.74; 95% CI 0.68–0.81). By contrast, Subtype B followed the classical undernutrition gradient: wealth quintile declined monotonically to the Richest (0.25; 95% CI 0.22–0.28), and maternal education declined monotonically to Higher (0.49; 95% CI 0.42–0.56). Ten of 17 covariates showed opposite effect directions between Subtype A and Subtype B (Table 1). The country-level intra-class correlation coefficient was 0.020, indicating that between-country variation accounted for approximately 2% of total variance after adjustment.

**Table 1.** Adjusted odds ratios from multilevel logistic regression for Subtype A (overweight mother with stunted or wasted child) and Subtype B (underweight mother with stunted or wasted child). Reference categories are noted below. All associations are significant at p < 0.001 unless otherwise indicated. Ten of 17 covariates show opposite effect directions (marked *).

| Covariate | Subtype A — OR (95% CI) | Subtype B — OR (95% CI) |  |
| --- | --- | --- | --- |
| *Maternal age (ref: 25–34 years)* |  |  |  |
| 15–24 | 0.67 (0.64–0.70) | 1.43 (1.35–1.51) | * |
| 35–49 | 1.33 (1.28–1.39) | 0.86 (0.80–0.92) | * |
| *Maternal education (ref: None)* |  |  |  |
| Primary | 1.23 (1.16–1.30) | 0.76 (0.70–0.81) | * |
| Secondary | 0.89 (0.83–0.94) | 0.71 (0.66–0.76) |  |
| Higher | 0.74 (0.68–0.81) | 0.49 (0.42–0.56) |  |
| *Wealth quintile (ref: Poorest)* |  |  |  |
| Poorer | 1.10 (1.04–1.17) | 0.72 (0.68–0.77) | * |
| Middle | 1.18 (1.12–1.25) | 0.52 (0.48–0.56) | * |
| Richer | 1.22 (1.14–1.30) | 0.40 (0.37–0.44) | * |
| Richest | 1.17 (1.09–1.26) | 0.25 (0.22–0.28) | * |
| *Residence (ref: Urban)* |  |  |  |
| Rural | 0.86 (0.82–0.91) | 1.07 (0.99–1.15) | * |
| *Child sex (ref: Male)* |  |  |  |
| Female | 0.88 (0.85–0.92) | 0.91 (0.86–0.95) |  |
| *Child age (ref: 24–35 months)* |  |  |  |
| 0–11 | 0.58 (0.55–0.62) | 0.75 (0.69–0.82) |  |
| 12–23 | 0.82 (0.78–0.87) | 1.33 (1.24–1.43) | * |
| 36–47 | 0.96 (0.91–1.01) | 0.98 (0.91–1.06) |  |

| Covariate | Subtype A — OR (95% CI) | Subtype B — OR (95% CI) |
| --- | --- | --- |
| 48–59 | 0.77 (0.73–0.81) | 0.87 (0.80–0.94) |
| *Country income group (ref: Low income)* |  |  |
| Lower-middle | 1.22 (0.97–1.52) | 1.26 (0.72–2.19) |
| Upper-middle | 1.98 (1.43–2.73) | 0.15 (0.06–0.34) * |

Note: Models adjusted for all listed covariates plus country random intercept. Intra-class correlation coefficient = 0.020. n = 181,629 dyads (seven excluded for covariate missingness). Asterisks denote opposite-direction effects between Subtype A and Subtype B, where at least one direction is statistically significant at $p < 0.05$ .

**Fig 2.**
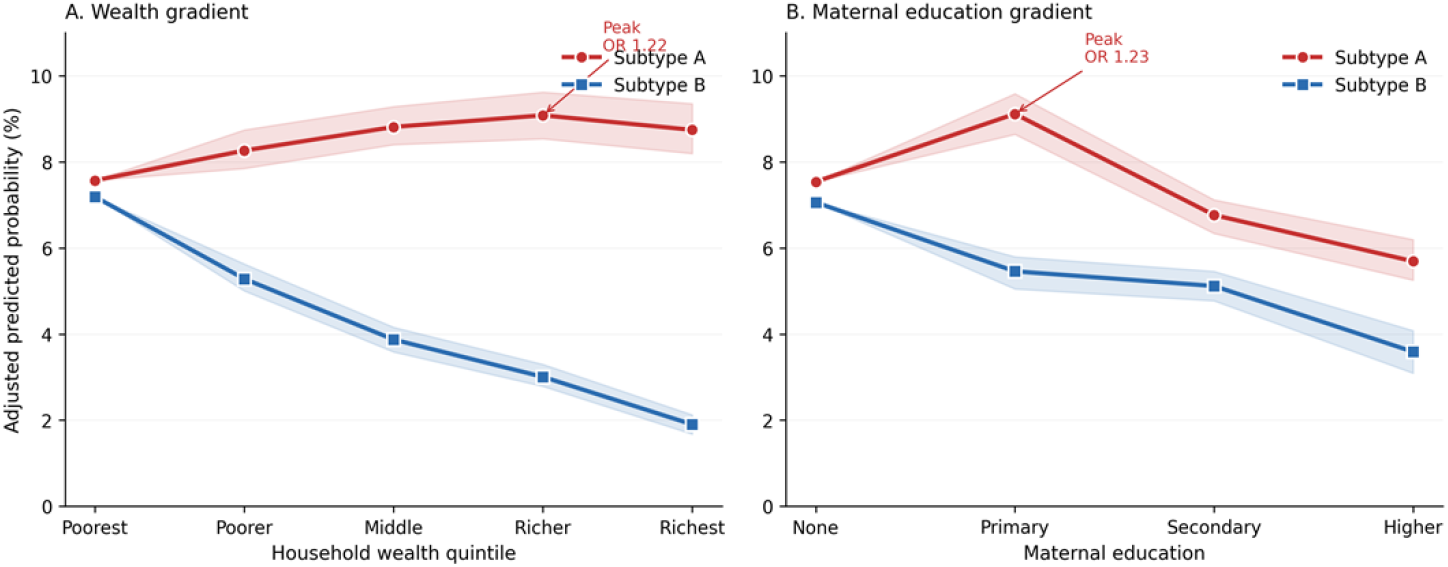
Adjusted predicted probabilities of Subtype A and Subtype B along wealth (Panel A) and maternal education (Panel B) gradients, derived from two-level random-intercept logistic models. Shaded ribbons are 95% Wald confidence intervals. Subtype A peaks at the Richer wealth quintile and at Primary maternal education (both annotated); Subtype B declines monotonically. The visualisation approach follows Seferidi and colleagues [9] and Chen and colleagues [8].

The Subtype A inverted-U adds new information to, and in part revises, Chen and colleagues [8]. Our monotonic Subtype B model recovers their 45-country finding that higher maternal education predicts lower overall DBM; our inverted-U Subtype A model shows that for this subtype the highest risk falls not at the socioeconomic summit but in the transitional middle. This non-monotonic pattern is consistent with the long-standing obesity–socioeconomic-status literature in LMICs [10–12] and with the Popkin nutrition-transition framework [2,3].

### 3.3. Machine-learning discrimination is moderate and signals absence of anthropometric shortcuts

The label-leakage audit confirmed that anthropometric variables alone were near-perfectly informative of all three outcomes (AUC > 0.99 for Subtype A, Subtype B, and any-DBM), justifying their exclusion from the feature set. In held-out leave-one-country-out cross-validation, xgboost achieved a mean area under the curve of 0.615 for Subtype A (range 0.539–0.662 across 30 countries) and 0.652 for Subtype B (range 0.562–0.762) (S1 Fig). Discrimination was lowest for Subtype A in Honduras (0.539), Tajikistan (0.569), and Pakistan (0.580)—countries whose unweighted country-specific gradients were flat or inverted—and highest in Kenya (0.762) and Cameroon (0.758) for Subtype B. These values indicate moderate discrimination from non-anthropometric survey variables alone, consistent with other LMIC social-determinant prediction models in the literature [16,28].

### 3.4. ML targeting doubles Subtype A recall but only modestly improves Subtype B recall

Targeting performance differed markedly between subtypes (Fig 3; Table 2). At a 20% budget— a realistic constrained-coverage operating point for an LMIC nutrition programme—ML-based targeting captured 35.3% of Subtype A cases (95% CI 29.1–39.8) versus 18.4% (95% CI 16.5– 21.7) for education-based targeting, a relative gain of 92%. Strikingly, education-based targeting was statistically indistinguishable from random (19.3%; 95% CI 18.5–20.0), showing little discriminatory value for Subtype A at this budget threshold. For Subtype B, ML captured 37.5% (95% CI 31.7–47.7) versus 32.1% (95% CI 26.3–42.2) for education-based and 20.0% (95% CI 18.6–21.0) for random—a 17% relative ML gain, substantial but far smaller than for Subtype A, and reflecting the fact that for monotonic gradients maternal education already serves as a reasonable proxy [8]. The asymmetry was robust across the 10% and 30% budgets.

**Table 2.** Targeting performance at the 20% budget threshold: weighted recall and precision for three targeting rules across two DBM subtypes. Confidence intervals from country-clustered bootstrap with 500 resamples.

| Subtype | Targeting rule | Recall — % (95% CI) | Precision — % (95% CI) |
| --- | --- | --- | --- |
| **Subtype A** | Random | 19.3 (18.5, 20.0) | 7.9 (6.7, 9.7) |
|  | Education-based | 18.4 (16.5, 21.7) | 7.6 (6.0, 10.3) |
|  | **ML-based** | **35.3 (29.1, 39.8)** | **14.5 (10.5, 19.2)** |
| **Subtype B** | Random | 20.0 (18.6, 21.0) | 4.3 (2.3, 6.6) |
|  | Education-based | 32.1 (26.3, 42.2) | 6.9 (4.3, 9.5) |
|  | **ML-based** | **37.5 (31.7, 47.7)** | **8.1 (5.1, 11.0)** |

Note: ML-based rule for Subtype A achieves a 92% relative gain in recall over education-based; for Subtype B the corresponding gain is 17%.

**Table 3.**
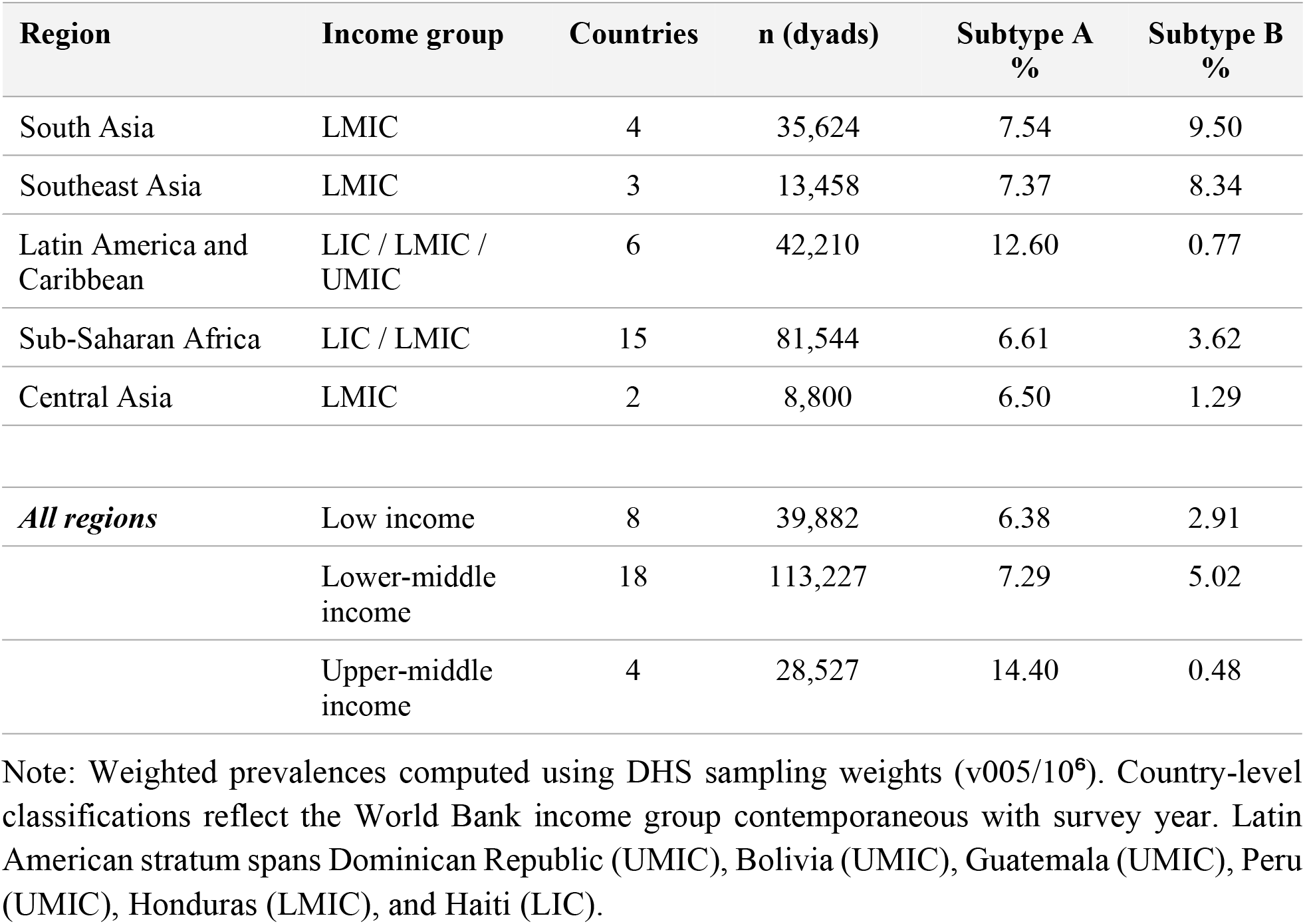
Weighted prevalence of concurrent maternal–child DBM subtypes by region and World Bank income group. Stratification approach follows Seferidi et al. (2022) and Popkin et al. (2020). n refers to total mother–child dyads in the stratum.

**Fig 3.**
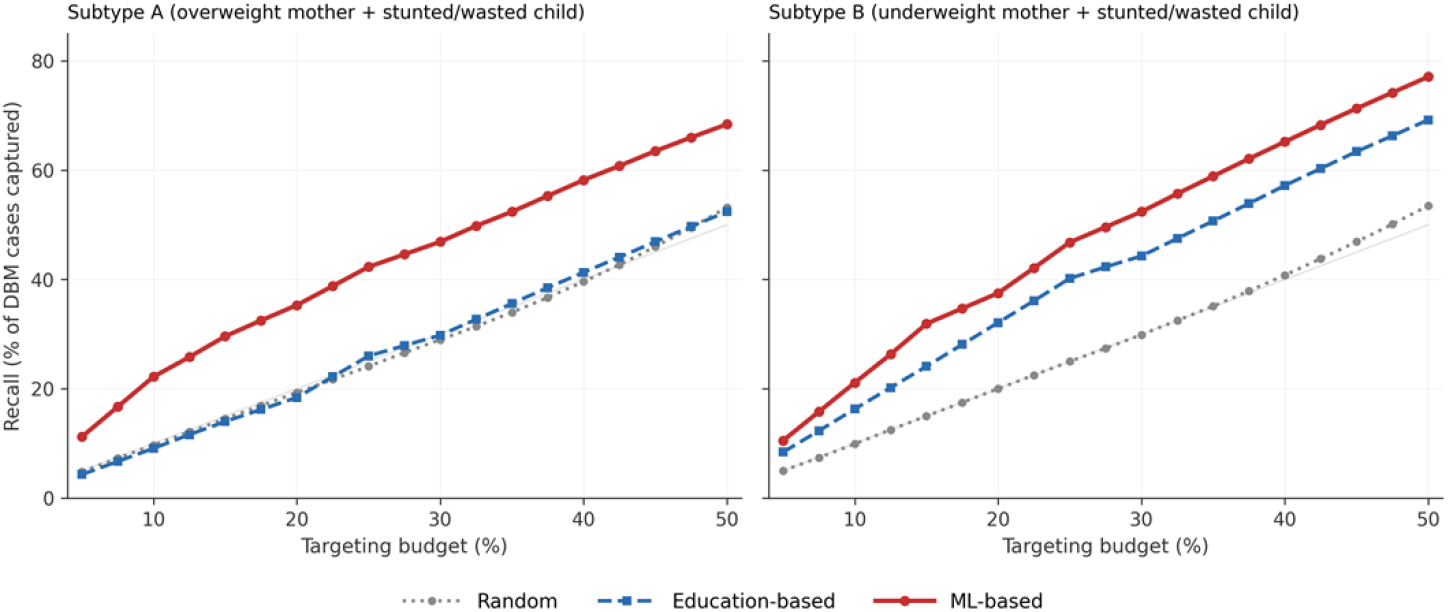
Recall-versus-budget curves for three targeting rules (random, education-based, and ML-based) for Subtype A and Subtype B, based on DHS Phase 7–8 pooled data from 30 LMICs. Held-out predictions from leave-one-country-out cross-validation. Budgets represent the fraction of the study population that could be reached by a nutrition-targeting programme; recall is the fraction of true DBM cases captured.

Decomposing ML gains by wealth-by-residence strata revealed that the two subtypes exhibited fundamentally different redistributive logics. For Subtype A, every stratum showed a positive net capture under ML relative to education-based targeting: the smallest gain was in the Poorest-urban cell (+22 additional cases captured) and the largest in the Richer-urban cell (+460), indicating that ML is universally better but disproportionately more informative where education is non-discriminating. For Subtype B, by contrast, middle- and upper-wealth cells showed net losses under ML (Middle-rural −95, Middle-urban −47, Richer-rural −36, Richer-urban −23, Richest-urban −19), while very large net gains concentrated in the Poorest-rural cell (+534) and the Poorer-rural cell (+169). Relative to a low-education-first rule, ML substantially reallocates Subtype B recall from middle-wealth strata to the poorest rural dyads—zero-sum along the wealth gradient rather than universally improving.

### 3.5. Fairness gaps on policy-relevant axes are concentrated in high-resource strata

The fairness audit at the 20% budget (Fig 4) revealed substantial group-level disparities that would remain invisible in an aggregate-accuracy framing. Policy-relevant axes—country income, maternal education, wealth, and maternal age—exhibited large equalized-odds and demographic-parity gaps; calibration gaps were uniformly small; and child-sex gaps were small throughout. For Subtype A, the largest equalized-odds gaps were on country income (0.570), maternal education (0.355), and maternal age (0.314). For Subtype B, they were on maternal education (0.591), wealth (0.559), and country income (0.426). Calibration gaps stayed below 0.006 on all 12 subtype-by-axis combinations, indicating that the predicted probabilities were well aligned with observed prevalence within groups; what differs across groups is not how well-calibrated the probabilities are but how much probability mass gets allocated to each group, and therefore how many group members clear any fixed inclusion threshold. Child-sex gaps reached at most 0.106 for Subtype A and 0.032 for Subtype B, consistent with child sex being largely uninformative for DBM risk in this analytic pool.

**Fig 4.**
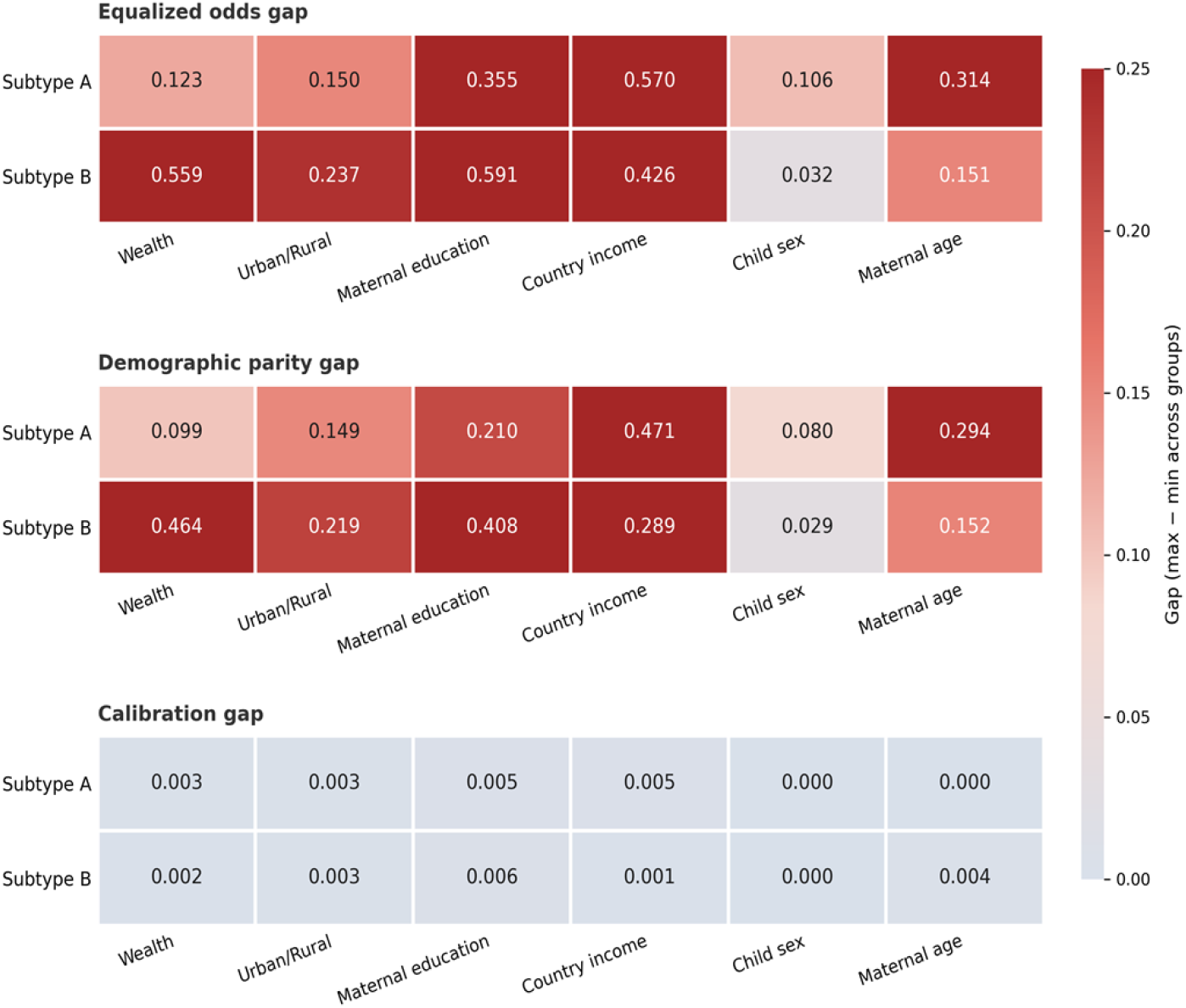
Fairness audit of ML-based targeting across six social axes at a 20% targeting budget. Cells show the maximum–minimum gap across within-axis groups for equalized-odds, demographic-parity, and calibration metrics. Warmer colours indicate larger group-level disparities.

A closer look at who the algorithm most often missed produced a counter-intuitive pattern. On every axis, the worst-off group—the group with the lowest true-positive rate—was a high-resource group. The Richest wealth quintile was worst-off for both subtypes (Subtype A true-positive rate 0.269; Subtype B 0.038). Higher-educated mothers were the worst-off education group (Subtype A 0.165; Subtype B 0.046). For Subtype A, low-income countries were worst-off (0.167). For Subtype B, urban residents were captured less efficiently than rural ones (urban true-positive rate 0.182). We return to the mechanism behind this pattern in Section 4.3.

### 3.6. Sensitivity to India down-sampling

The full-India sensitivity analysis (354,691 dyads; 55.8% Indian) reproduced the main-analysis performance nearly exactly. Country-specific AUC differences between main and sensitivity models had a mean absolute value below 0.01 for both subtypes (Subtype A mean main AUC 0.615 versus sensitivity 0.605; Subtype B 0.652 versus 0.644). For 83.3% of countries in the Subtype A analysis and 70.0% in the Subtype B analysis, |ΔAUC| < 0.02, confirming that PSU-level down-sampling of India did not systematically distort predictive performance.

## 4. Discussion

### 4.1. Principal findings

Two findings from this study matter most. The first is substantive: Subtype A and Subtype B are not socially distributed in the same way. They follow contrasting gradients—inverted-U versus monotonic—and disagree on the direction of 10 of the 17 covariates we examined, a heterogeneity that pooled analyses cannot capture. The second is operational: the value of ML depends heavily on which subtype is being targeted. At a 20% budget, ML lifted Subtype A recall by 92% over an education-based rule—because the education rule was no better than random for Subtype A—but improved Subtype B recall by only 17%, and did so largely by reallocating capture from middle-to low-wealth strata. We also found that the groups most often missed were not poor households, but the rare positives in high-resource strata. To our knowledge, no prior study has examined concurrent maternal–child DBM through this combined lens.

### 4.2. Comparison with prior literature

Two prior bodies of work frame our results most directly. Chen and colleagues [8] showed that the direction of the maternal-education effect on DBM depends on which subtype is considered, and our monotonic Subtype B model replicates their finding (see also [39]). What our data add is that for Subtype A the gradient is not merely positive but non-linear, with the highest risk falling in the middle of both the wealth and education distributions—a pattern the nutrition-transition literature [2,10–12] has long predicted but which pooled multi-country models rarely localise to a specific peak. This middle-peak specification matters for programme design, because regional studies of similar subtypes in sub-Saharan Africa [14] and South and Southeast Asia [13] have so far reported pooled rather than stratified gradients. On the fairness side, our audit extends a growing literature on algorithmic fairness in health [20,24,25,40] from high-income clinical settings to LMIC nutrition targeting; we are not aware of a prior fairness audit for this outcome. The cross-level framework by Eom and colleagues [41] examined child anthropometric outcomes in 28 sub-Saharan African countries with women’s empowerment as the principal exposure; our work is complementary in bringing algorithmic fairness—rather than the presence of inequality alone—into the analytic foreground.

### 4.3. Why algorithmic missingness concentrates in high-resource strata

This is not the pattern reported by Obermeyer and colleagues [20]. In their case a biased proxy label—healthcare cost standing in for medical need—produced racially disparate access. Our label, by contrast, is the ground-truth anthropometric outcome itself, and the gap has a more ordinary explanation. Subtype B prevalence in the Richest wealth quintile is roughly 1.5%; in the Poorest quintile it exceeds 7%. A probability-ranked classifier constrained to a 20% budget will therefore draw almost entirely from the higher-prevalence strata, and the few true positives in the low-prevalence tail stay unselected. The wealth and residence axes behave the same way for Subtype B: the small number of positives in the Richest quintile, and in urban areas, get crowded out by the much larger number in lower-wealth and rural strata. This is essentially the calibration-versus-balance trade-off formalised by Kleinberg and colleagues [23] and Chouldechova [22], showing up in a targeting decision. In practice, a programme using an off-the-shelf ML eligibility rule to reach Subtype B among the poor is making a statistically correct bet; but if the goal is universal coverage of high-risk households regardless of how common risk is in each stratum, a single pooled rule is the wrong tool. Stratified budgets—one allocation per residence or wealth group— or group-specific thresholds fit that goal better.

### 4.4. Implications for nutrition-targeting policy

The policy implication is straightforward. A single targeting rule for both subtypes is likely to be wasteful: a maternal-education threshold is close to random for Subtype A and a reasonable proxy for Subtype B. If a programme wants to detect the transitional Subtype A cases, it needs a rule that is sensitive to the middle of the wealth and education distributions—the main case for using ML. For Subtype B the comparison is more subtle. ML and an education rule achieve similar overall recall but reach different households: ML moves capture toward the rural poor and away from middle-wealth strata. That is a choice about who gets reached, not an accuracy upgrade, and programmes should make it explicitly before the algorithm is trained rather than after. A parallel debate about how targeting redistributes benefits has long been active in the cash-transfer literature [17–19]. The interventions themselves are not the main bottleneck; the evidence-based nutrition toolkit is well established [7,36,42]. What has been missing is the decision rule that gets interventions to the right households—and then a country-by-country check that the chosen rule does not systematically leave some groups behind [26,27,43].

### 4.5. Strengths and limitations

This study pooled 181,636 dyads from 30 LMICs under standardised DHS protocols [29,30,32], used strict label-leakage defences, and validated the model with leave-one-country-out cross-validation, which simulates the most realistic deployment scenario—transfer to a new country. Country-clustered bootstrap inference accounted for between-country heterogeneity, and a full-India sensitivity analysis confirmed that PSU down-sampling did not distort either discrimination or fairness patterns.

The limitations that matter most for this paper are the ones specific to it. First, DHS data are cross-sectional, so the Subtype A inverted-U cannot be read as a developmental trajectory; the Popkin-style nutrition-transition interpretation is compatible with our data but not established by them. Second, the moderate LOCO-CV AUC (0.615–0.652) reflects a deliberate choice to use only DHS questionnaire variables; performance may be higher when phone data, satellite imagery, or biomarkers are also available [16,17]. Third, a pooled multi-country model trades country-specific accuracy for transferability, and the large country-income and education fairness gaps for Subtype A suggest that country-specific recalibration is likely to be necessary before deployment. Fourth, six countries were excluded because maternal BMI or child anthropometric variables were absent from the relevant public recodes, which may bias prevalence estimates toward countries with stronger measurement infrastructure. Fifth, fairness was audited post hoc; whether fairness-aware training [40] could recover the missed high-resource Subtype A cases without substantial recall loss is a question we leave for future work. Finally, we focused on the two within-dyad concurrent subtypes defined by the Lancet series [2,3] and did not examine maternal overweight paired with child overweight, which is a distinct phenomenon more salient in upper-middle-income settings.

### 4.6. Conclusions

Concurrent maternal–child DBM in LMICs behaves like two problems, not one. Subtype A and Subtype B follow different social gradients, and a targeting rule that works for one of them may be close to useless for the other. ML substantially improves the identification of Subtype A, where the common education proxy is no better than random; for Subtype B the gain is smaller, and it comes from reallocating capture from middle-to low-wealth households rather than from a clear accuracy advantage. The households the algorithm most often misses are not the poor: they are the rare positives in the highest-wealth and highest-education groups, where the base rate is simply too low for a fixed-budget rule to reach. Before using such a rule, programmes should decide whether their priority is total capture or the distribution of capture—and audit accordingly. That decision is a policy choice, not an algorithmic one.

## Author contributions

Conceptualization: Xiaoyu Wu. Methodology: Xiaoyu Wu, Benqing Zheng. Software: Xiaoyu Wu, Benqing Zheng. Formal analysis: Xiaoyu Wu, Benqing Zheng. Data curation: Xiaoyu Wu. Validation: Benqing Zheng. Visualization: Xiaoyu Wu. Writing – original draft: Xiaoyu Wu. Writing – review & editing: Xiaoyu Wu, Benqing Zheng.

## Data availability

All DHS data underlying the findings are publicly available to registered researchers from The DHS Program (https://dhsprogram.com). The UNICEF/WHO/World Bank Group Joint Child Malnutrition Estimates 2025 edition is available from https://data.unicef.org/resources/jme-report-2025/. All analysis code is permanently deposited at Zenodo (concept DOI: https://doi.org/10.5281/zenodo.19731115; this version: https://doi.org/10.5281/zenodo.19731116) under the MIT License, and is accompanied by the STROBE checklist and the TRIPOD+AI checklist. The deposited pipeline reproduces every number, table, and figure reported in this manuscript.

## Declaration of generative AI use in the writing process

During the preparation of this manuscript, the authors used generative AI tools for language editing and to support reference cross-checking. After using these tools, the authors reviewed and edited the content as needed and take full responsibility for the content of the publication.

## Supporting information

**S1 Fig.**
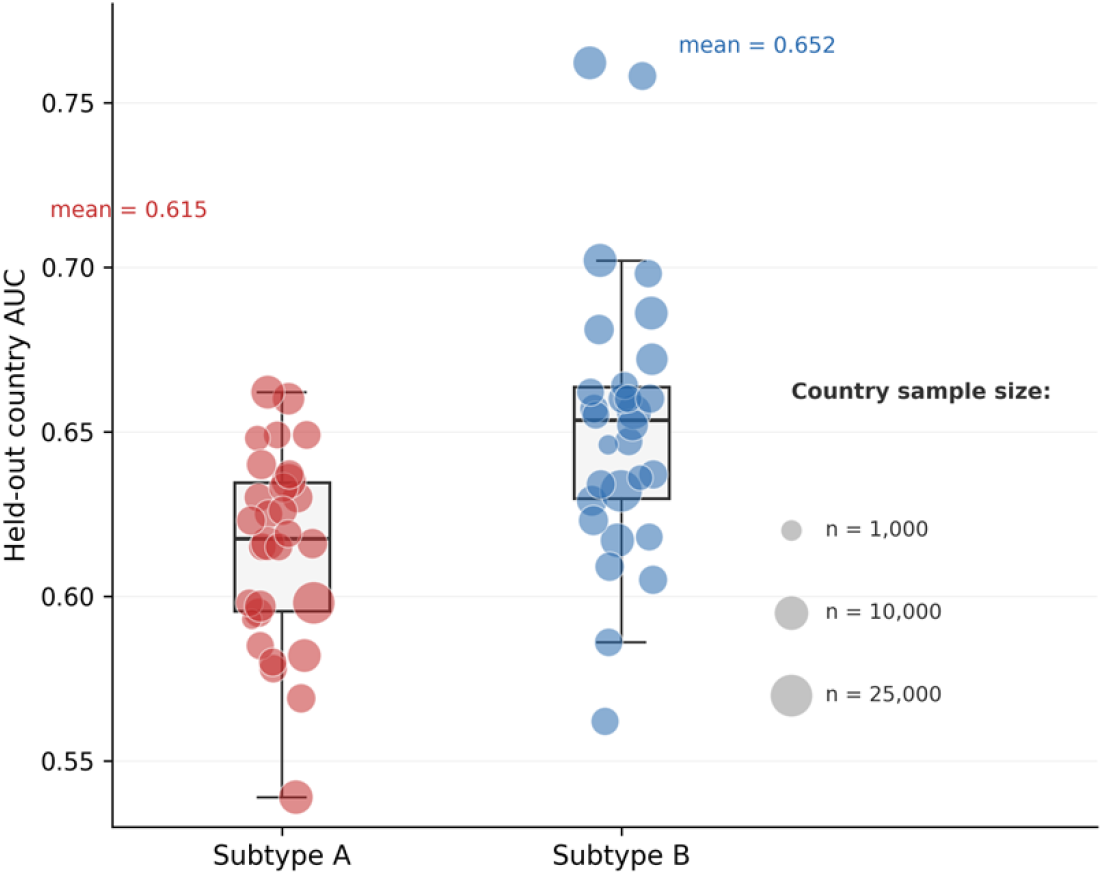
Leave-one-country-out cross-validation area-under-curve distribution for Subtype A and Subtype B. Each point represents one held-out country (30 folds per outcome); marker size is proportional to country sample size. Boxes show the interquartile range; horizontal lines show the median.

## References

1. World Health Organization, United Nations Children’s Fund (UNICEF), World Bank Group. Levels and trends in child malnutrition: UNICEF / WHO / World Bank Group joint child malnutrition estimates — key findings of the 2025 edition. Geneva: World Health Organization; 2025. Licence: CC BY-NC-SA 3.0 IGO.

2. Popkin BM, Corvalan C, Grummer-Strawn LM. Dynamics of the double burden of malnutrition and the changing nutrition reality. Lancet. 2020;395(10217):65–74. doi:10.1016/S0140-6736(19)32497-3

3. Wells JC, Sawaya AL, Wibaek R, Mwangome M, Poullas MS, Yajnik CS, et al. The double burden of malnutrition: aetiological pathways and consequences for health. Lancet. 2020;395(10217):75–88. doi:10.1016/S0140-6736(19)32472-9

4. Caballero B. A nutrition paradox — underweight and obesity in developing countries. N Engl J Med. 2005;352(15):1514–1516. doi:10.1056/NEJMp048310

5. Victora CG, Christian P, Vidaletti LP, Gatica-Domínguez G, Menon P, Black RE. Revisiting maternal and child undernutrition in low-income and middle-income countries: variable progress towards an unfinished agenda. Lancet. 2021;397(10282):1388–1399. doi:10.1016/S0140-6736(21)00394-9

6. Nugent R, Levin C, Hale J, Hutchinson B. Economic effects of the double burden of malnutrition. Lancet. 2020;395(10218):156–164. doi:10.1016/S0140-6736(19)32473-0

7. Escher NA, Andrade GC, Ghosh-Jerath S, Millett C, Seferidi P. The effect of nutrition-specific and nutrition-sensitive interventions on the double burden of malnutrition in low-income and middle-income countries: a systematic review. Lancet Glob Health. 2024;12(3):e419–e432. doi:10.1016/S2214-109X(23)00562-4

8. Chen S, Richardson S, Kong Y, Ma N, Zhao A, Song Y, et al. Association between parental education and simultaneous malnutrition among parents and children in 45 low- and middle-income countries. JAMA Netw Open. 2023;6(1):e2251727. doi:10.1001/jamanetworkopen.2022.51727

9. Seferidi P, Hone T, Duran AC, Bernabe-Ortiz A, Millett C. Global inequalities in the double burden of malnutrition and associations with globalisation: a multilevel analysis of Demographic and Health Surveys from 55 low-income and middle-income countries, 1992–2018. Lancet Glob Health. 2022;10(4):e482–e490. doi:10.1016/S2214-109X(21)00594-5

10. Monteiro CA, Moura EC, Conde WL, Popkin BM. Socioeconomic status and obesity in adult populations of developing countries: a review. Bull World Health Organ. 2004;82(12):940–946.

11. Dinsa GD, Goryakin Y, Fumagalli E, Suhrcke M. Obesity and socioeconomic status in developing countries: a systematic review. Obes Rev. 2012;13(11):1067–1079. doi:10.1111/j.1467-789X.2012.01017.x

12. Jaacks LM, Vandevijvere S, Pan A, McGowan CJ, Wallace C, Imamura F, et al. The obesity transition: stages of the global epidemic. Lancet Diabetes Endocrinol. 2019;7(3):231–240. doi:10.1016/S2213-8587(19)30026-9

13. Talukder A, Kelly M, Gray D, Sarma H. Influence of maternal education and household wealth on double burden of malnutrition in South and Southeast Asia. Matern Child Nutr. 2025;21(4):e70049. doi:10.1111/mcn.70049

14. Bawuah A, Baatiema L, Sarfo M, Appiah F, Yaya S. Malnourished child, overweight mother? Examining the double burden of malnutrition in sub-Saharan Africa. Matern Child Nutr. 2026;22:e70175. doi:10.1111/mcn.70175

15. Ijaiya MA, Anjorin S, Uthman OA. Income and education disparities in childhood malnutrition: a multi-country decomposition analysis. BMC Public Health. 2024;24:2882. doi:10.1186/s12889-024-20378-z

16. Aiken E, Bellue S, Karlan D, Udry C, Blumenstock JE. Machine learning and phone data can improve targeting of humanitarian aid. Nature. 2022;603(7903):864–870. doi:10.1038/s41586-022-04484-9

17. Smythe IS, Blumenstock JE. Geographic microtargeting of social assistance with high-resolution poverty maps. Proc Natl Acad Sci USA. 2022;119(32):e2120025119. doi:10.1073/pnas.2120025119

18. Brown C, Ravallion M, van de Walle D. A poor means test? Econometric targeting in Africa. J Dev Econ. 2018;134:109–124. doi:10.1016/j.jdeveco.2018.05.004

19. Hanna R, Olken BA. Universal basic incomes versus targeted transfers: anti-poverty programs in developing countries. J Econ Perspect. 2018;32(4):201–226. doi:10.1257/jep.32.4.201

20. Obermeyer Z, Powers B, Vogeli C, Mullainathan S. Dissecting racial bias in an algorithm used to manage the health of populations. Science. 2019;366(6464):447–453. doi:10.1126/science.aax2342

21. Hardt M, Price E, Srebro N. Equality of opportunity in supervised learning. In: Advances in Neural Information Processing Systems 29. 2016. p. 3315–3323.

22. Chouldechova A. Fair prediction with disparate impact: a study of bias in recidivism prediction instruments. Big Data. 2017;5(2):153–163. doi:10.1089/big.2016.0047

23. Kleinberg J, Mullainathan S, Raghavan M. Inherent trade-offs in the fair determination of risk scores. In: Proceedings of the 8th Innovations in Theoretical Computer Science Conference (ITCS 2017). LIPIcs 67, 43:1–43:23. doi:10.4230/LIPIcs.ITCS.2017.43

24. Rajkomar A, Hardt M, Howell MD, Corrado G, Chin MH. Ensuring fairness in machine learning to advance health equity. Ann Intern Med. 2018;169(12):866–872. doi:10.7326/M18-1990

25. Mhasawade V, Zhao Y, Chunara R. Machine learning and algorithmic fairness in public and population health. Nat Mach Intell. 2021;3(8):659–666. doi:10.1038/s42256-021-00373-4

26. Collins GS, Moons KGM, Dhiman P, Riley RD, Beam AL, Van Calster B, et al. TRIPOD+AI statement: updated guidance for reporting clinical prediction models that use regression or machine learning methods. BMJ. 2024;385:e078378. doi:10.1136/bmj-2023-078378

27. World Health Organization. Ethics and governance of artificial intelligence for health: WHO guidance. Geneva: World Health Organization; 2021. Licence: CC BY-NC-SA 3.0 IGO.

28. Rao B, Rashid M, Hasan MG, Thunga G. Machine learning in predicting child malnutrition: a meta-analysis of Demographic and Health Surveys data. Int J Environ Res Public Health. 2025;22(3):449. doi:10.3390/ijerph22030449

29. Corsi DJ, Neuman M, Finlay JE, Subramanian SV. Demographic and health surveys: a profile. Int J Epidemiol. 2012;41(6):1602–1613. doi:10.1093/ije/dys184

30. Croft TN, Marshall AMJ, Allen CK, et al. Guide to DHS Statistics (DHS-7). Rockville, Maryland: ICF; 2018.

31. WHO Multicentre Growth Reference Study Group. WHO Child Growth Standards: Length/Height-for-Age, Weight-for-Age, Weight-for-Length, Weight-for-Height and Body Mass Index-for-Age: Methods and Development. Geneva: World Health Organization; 2006.

32. Assaf S, Kothari MT, Pullum TW. An assessment of the quality of DHS anthropometric data, 2005– 2014. DHS Methodological Reports No. 16. Rockville, Maryland: ICF International; 2015.

33. Rutstein SO, Johnson K. The DHS Wealth Index. DHS Comparative Reports No. 6. Calverton, Maryland: ORC Macro; 2004.

34. Howe LD, Galobardes B, Matijasevich A, Gordon D, Johnston D, Onwujekwe O, et al. Measuring socio-economic position for epidemiological studies in low-and middle-income countries: a methods of measurement in epidemiology paper. Int J Epidemiol. 2012;41(3):871–886. doi:10.1093/ije/dys037

35. Chen T, Guestrin C. XGBoost: a scalable tree boosting system. In: Proceedings of the 22nd ACM SIGKDD International Conference on Knowledge Discovery and Data Mining. New York: ACM; 2016. p. 785–794. doi:10.1145/2939672.2939785

36. Bhutta ZA, Das JK, Rizvi A, Gaffey MF, Walker N, Horton S, et al. Evidence-based interventions for improvement of maternal and child nutrition: what can be done and at what cost? Lancet. 2013;382(9890):452–477. doi:10.1016/S0140-6736(13)60996-4

37. von Elm E, Altman DG, Egger M, Pocock SJ, Gøtzsche PC, Vandenbroucke JP. The Strengthening the Reporting of Observational Studies in Epidemiology (STROBE) statement: guidelines for reporting observational studies. Lancet. 2007;370(9596):1453–1457. doi:10.1016/S0140-6736(07)61602-X

38. Benjamini Y, Hochberg Y. Controlling the false discovery rate: a practical and powerful approach to multiple testing. J R Stat Soc Series B Methodological. 1995;57(1):289–300. doi:10.1111/j.2517-6161.1995.tb02031.x

39. Li Z, Kim R, Vollmer S, Subramanian SV. Factors associated with child stunting, wasting, and underweight in 35 low- and middle-income countries. JAMA Netw Open. 2020;3(4):e203386. doi:10.1001/jamanetworkopen.2020.3386

40. Pierson E, Cutler DM, Leskovec J, Mullainathan S, Obermeyer Z. An algorithmic approach to reducing unexplained pain disparities in underserved populations. Nat Med. 2021;27(1):136–140. doi:10.1038/s41591-020-01192-7

41. Eom YJ, Chi H, Jung S, Kim J, Jeong J, Subramanian SV, et al. Women’s empowerment and child anthropometric failures across 28 sub-Saharan African countries: a cross-level interaction by Gender Inequality Index. SSM Popul Health. 2024;26:101651. doi:10.1016/j.ssmph.2024.101651

42. Hawkes C, Ruel MT, Salm L, Sinclair B, Branca F. Double-duty actions: seizing programme and policy opportunities to address malnutrition in all its forms. Lancet. 2020;395(10218):142–155. doi:10.1016/S0140-6736(19)32506-1

43. Constenla-Villoslada S, Liu Y, McBride L, Ouma C, Mutanda N, Barrett CB. High-frequency monitoring enables machine learning–based forecasting of acute child malnutrition for early warning. Proc Natl Acad Sci USA. 2025;122(23):e2416161122. doi:10.1073/pnas.2416161122

